# Dedifferentiation of memory-related brain function as a potential early biomarker of Alzheimer’s disease in asymptomatic older women: Results from the PREVENT-AD Cohort

**DOI:** 10.1101/2020.11.30.20241117

**Authors:** Sheida Rabipour, Sricharana Rajagopal, Stamatoula Pasvanis, PREVENT-AD Research Group, M. Natasha Rajah

**Affiliations:** Centre for Cerebral Imaging, Douglas Hospital Research Centre, Montreal, Canada; Department of Psychiatry, McGill University, Montreal, Canada

**Author notes:** Corresponding Author: M. Natasha Rajah, Ph.D. Room, 2114, CIC Pavilion, Douglas Hospital Research Centre, Verdun, QC, H4H 1R3. Data used in preparation of this article were obtained from the Pre-symptomatic Evaluation of Novel or Experimental Treatments for Alzheimer’s Disease (PREVENT-AD) program (https://douglas.research.mcgill.ca/stop-ad-centre), data release 5.0 (November 30, 2017). A complete listing of PREVENT-AD Research Group can be found in the PREVENT-AD database: https://preventad.loris.ca/acknowledgements/acknowledgements.php?date=[2019-06-03]. The investigators of the PREVENT-AD program contributed to the design and implementation of PREVENT-AD and/or provided data but did not participate in analysis or writing of this report.

**Keywords:** Apolipoprotein E ε4 polymorphism, Alzheimer’s disease, Associative learning, Brain-behavior relationships, Dedifferentiation, Episodic memory, Familial history, Partial least squares analysis, Sex differences, Task-related functional MRI

## Abstract

Late-onset Alzheimer’s disease (AD) disproportionately affects women compared to men. Episodic memory decline is one of the earliest and most pronounced deficits observed in AD. However, it remains unclear whether there are sex differences in episodic memory-related brain function in cognitively intact older adults at risk of developing AD. In the current study, we used task fMRI to test for sex differences in episodic memory-related brain activity and brain activity-behavior correlations in cognitively intact older adults with a family history of AD from the PREVENT-AD cohort study in Montreal, Canada (M_age_=63.03±3.78; M_education_=15.41±3.40). Importantly, we tested women and men who were matched in age, body mass index, years of education, and proximity to the age of parental AD onset. We used data-driven task-based multivariate partial least squares (PLS) analysis to identify sex differences in brain activity during the successful encoding and retrieval of objects and their associated spatial context. We used behavior PLS to examine sex differences in the correlations between brain activity and memory performance at encoding and retrieval. Our results suggested no significant sex differences in behavioral performance on the memory task. Yet, we observed sex differences in task-related brain activity and in brain activity-behavior correlations during the encoding of object-location associative memories and object-only item memory, and the retrieval of object only item memories. Specifically, subsequent object-location associative retrieval associated with encoding related activation of caudate, cingulate, and middle occipital cortex in women and, additionally, of temporo-parietal regions in men. We also found male-specific activations in ventrolateral prefrontal cortex (PFC) and insula during associated with the encoding and retrieval of object-only information. Moreover, whereas activity in ventrolateral PFC, posterior cingulate, and inferior parietal regions during encoding supported subsequent performance in women, activity in these regions during retrieval supported object-location associative memory in men. Similarly, we found that activity in ventrolateral PFC, precuneus, parahippocampal, anterior cingulate, and occipital regions during retrieval supported general memory performance in women but object-only retrieval in men. Our findings suggest functional dedifferentiation of episodic memory-related brain activation and performance in women compared to men. Follow up analyses should test for sex differences in the relationship between brain activity patterns and performance longitudinally, in association with risk factors for AD development.

## 1. Introduction

Late-onset sporadic Alzheimer’s Disease (AD) accounts for an estimated 70% of dementia cases worldwide and disproportionately affects women compared to men (World Health Organization, 2019). In the United States, 65% of AD cases are in women (Alzheimer’s Association, 2020; World Health Organization, 2019). Compared to men, women demonstrate elevated incidence and lifelong risk of AD even after accounting for sex and/or gender (sex/gender) differences in life expectancy, and exhibit more rapid cognitive decline and brain atrophy in the presence of AD-related neuropathology (Gamache, Yun, & Chiba-Falek, 2020). Despite advances in our overall understanding of AD neuropathophysiology, little is known about why there is this sex/gender difference in AD prevalence (Nebel et al., 2018).

One of the earliest and most pronounced deficits observed in AD is episodic memory decline (Buckner, 2004; McKhann et al., 2011). Behavioral studies have reported that females perform better than males on episodic memory tasks for verbal stimuli (Bleecker, Bolla-Wilson, Agnew, & Meyers, 1988; Bremner et al., 2001; Herlitz, Nilsson, & Backman, 1997; Kimura & Harshman, 1984; Ragland, Coleman, Gur, Glahn, & Gur, 2000), negative emotional stimuli (Young, Bellgowan, Bodurka, & Drevets, 2013), face stimuli (Keightley, Winocur, Burianova, Hongwanishkul, & Grady, 2006; W. Sommer, Hildebrandt, Kunina-Habenicht, Schacht, & Wilhelm, 2013; Yonker, Eriksson, Nilsson, & Herlitz, 2003) and verbal paired associative memory (Bender, Naveh-Benjamin, & Raz, 2010). In contrast, males appear to perform better than females on spatial navigation (Astur, Purton, Zaniewski, Cimadevilla, & Markus, 2016; Astur, Tropp, Sava, Constable, & Markus, 2004; Driscoll, Hamilton, Yeo, Brooks, & Sutherland, 2005) and object-location associative memory tasks (Postma, Izendoorn, & De Haan, 1998). Behavioral lifespan studies suggest these sex differences are stable across the adult lifespan (de Frias, Nilsson, & Herlitz, 2006; Jack et al., 2015). However, little research has explored whether and how sex/gender may interact with the effect of age and AD risk factors on episodic memory-related brain function – despite the; higher reported prevalence, incidence, and burden of AD in women compared to men (Beam et al., 2018; Mazure & Swendsen, 2016).

Prior studies provide evidence that sex/gender influences age-related differences in the neural correlates of episodic memory (Gur & Gur, 2002; McCarrey, An, Kitner-Triolo, Ferrucci, & Resnick, 2016; Subramaniapillai et al., 2019). These differences are even more pronounced in memory-related pathology: AD appears to impact memory functions more strongly and widely in women compared to men (Irvine, Laws, Gale, & Kondel, 2012). For example, although behavioral studies suggest that women score higher on verbal episodic memory tasks than men in normative aging and in clinical samples with preserved memory function, this pattern appears to shift in advanced stages of AD pathology, where women with AD tend to perform worse than men with AD (Brunet, Caldwell, Brandt, & Miller, 2020; Chapman et al., 2011). This effect may result from sex differences in the rate of clinical progression and brain atrophy in memory-related regions such as the medial temporal lobes, where women appear to decline more rapidly – independent of other risk factors (e.g., APOE4) – in healthy aging and at various stages of AD progression (Holland, Desikan, Dale, McEvoy, & Alzheimer’s Disease Neuroimaging, 2013; Lin et al., 2015). Similarly, hippocampal atrophy appears to progress more steeply in women, compared to men, in the presence of higher tau and lower amyloid-beta (Aβ) levels in the cerebrospinal fluid (Koran, Wagener, & Hohman, 2017). Such findings suggest a complex relationship between aging, sex, and AD-related neuropathology on memory.

To elucidate neural and behavioural processes underlying episodic memory changes related to healthy vs. pathological aging, studies increasingly focus on examining asymptomatic adults at higher risk of developing AD (Frankish & Horton, 2017). Such work suggests that early neural biomarkers or cognitive indices of AD development – e.g., *APOE4* carrier status, circulating tau and Aβ levels (Brookmeyer, Abdalla, Kawas, & Corrada, 2018; Kern et al., 2018; Leoutsakos, Gross, Jones, Albert, & Breitner, 2016; Molinuevo et al., 2016; Rabipour et al., 2020; Weiner & Veitch, 2015) – may exert different influences on neural processes in men and women (Buckley et al., 2020; Caldwell, Berg, Cummings, & Banks, 2017). For example, female *APOE4* carriers show greater brain hypometabolism and cortical thinning than male carriers, suggesting that women may be more susceptible to the metabolic effects of *APOE4* (Sampedro et al., 2015). Similarly, at low levels of Aβ and high levels of tau, women appear to have more rapid hippocampal atrophy as well as cognitive decline – a relationship that appears mediated, to some extent, by *APOE4* carrier status (Koran et al., 2017). Despite the influence of sex and gender on cognitive aging, relatively few studies have examined the effects of these factors on AD risk and development. Examining sex and gender in populations likely experiencing neurological changes associated with AD, before symptoms emerge, may help identify early markers or indices that contribute to differential AD development in men and women.

Because declines in episodic memory are among the earliest manifestations of AD (Bateman et al., 2012; Gardiner, 2001), tasks that differentially probe recognition and recall of contextual details associated with a past event are particularly well suited to detect early AD-related changes. For example, adults in prodromal states of AD, such as mild cognitive impairment, demonstrate poorer retrieval accuracy of object-location source associations as well as decreased volumes in AD-related brain regions such as medial temporal lobe, compared to controls (Hampstead, Towler, Stringer, & Sathian, 2018). Similarly, our previous work demonstrated different relationships between episodic memory performance and underlying neural processes in people at higher risk of AD, even when group differences in task performance are not observed (Rabipour et al., 2020; Rajah et al., 2017). Therefore, in the current study we use an object-location associative memory paradigm that allows us to differentiate brain activity during encoding and retrieval of objects and object-location association to determine if there are sex differences in episodic memory-related brain function in older adults with elevated AD risk, including family history of AD.

### The present Study

Here we evaluated sex/gender differences – via self-reported sex – in the behavioral and neural correlates of episodic memory in older adults who participated in the PREVENT-AD program (https://douglas.research.mcgill.ca/stop-ad-centre). Because our sample uniformly contained older adults with first-degree family history of AD, we aimed to examine the potential influence of self-reported sex on episodic memory performance and related brain activity over and above the influence of family history. Based on the prevalence of +*APOE4* genotype – 14% globally and 34-50% (Alzforum, 2010; Heffernan, Chidgey, Peng, Masters, & Roberts, 2016) in adults with a first degree relative with late-onset AD – we further sought to investigate whether *APOE4* carrier status would interact with sex on brain-behavior correlations.

Specifically, women and men were scanned during the encoding and retrieval phases of an object-location source memory paradigm. The experimental design was such that it allowed us to dissociated event-related activation related to successful encoding and retrieval of object-location associative memories (source hits) and object-only item memory (source failures). Multivariate task partial least squares (T-PLS) and behavior PLS (B-PLS) were used to test the hypotheses that: i) sex differences exist in mean levels of brain activity during successful encoding and retrieval of object-location spatial context associations (using T-PLS), and ii) sex differences exist in brain activity-memory performance correlations at encoding (subsequent memory effect) and retrieval of object-location association (source hits) (using B-PLS). We further explored whether any apparent sex differences in task-related activity or activity-performance correlations differed in older adults with an apolipoprotein E ε4 allele (+*APOE4*), compared to adults without this genetic risk for AD (-*APOE4*). The goal of this work is to help elucidate how AD risk differentially affects memory systems in women compared to men. Given the reportedly greater impact of AD on women compared to men (Gamache et al., 2020; Irvine et al., 2012), we hypothesized that women and men would exhibit distinct brain activation patterns related to the retrieval of object-location association, as well as different brain activity-behavior correlations during episodic retrieval. Importantly, in this study men and women were matched in age, body mass index (BMI), years of education, proximity to the age of parental AD onset (estimated years to AD onset; EYO), and motion during the fMRI scans, to help identify sex differences in brain activity and/or brain activity – memory performance correlations, after controlling for these confounds.

## 2. Methods

### 2.1 Participants: PREVENT-AD Cohort

Participants were recruited for the longitudinal PRe-symptomatic EValuation of Experimental or Novel Treatments for Alzheimer’s Disease (PREVENT-AD) program, an observational cohort study of asymptomatic older adults with first-degree family history of AD in Montreal, Canada (Breitner, Poirier, Etienne, & Leoutsakos, 2016). We evaluated baseline data in 88 age-and education-matched healthy older adults with elevated risk of AD due to family history who were enrolled up to August 31^st^, 2017 (i.e., data release 5.0) and participated in the task fMRI portion of the study (see below). From the full sample, we excluded participants based on confounding genetic factors (i.e., *APOE2* carriers, *n*=34; *APOE44* homozygotes, *n*=7; unavailable genotype, *n*=3); having below-chance performance or fewer than eight trials per response type in the task fMRI protocol (*n*= 95); and poor fMRI image resolution (*n*=32).

Because this sample was heavily unbalanced with respect to sex (43 men compared to 128 women), we selected a subset of 43 women matched in age, education, and *APOE4* carrier status to the original sample of men. After excluding age outliers, our final sample comprised 80 older adults (40 men and 40 women; M_age_=63.03±3.78; M_education_=15.41±3.40).

### 2.2 Protocol

Enrolment criteria for the PREVENT-AD trial (Breitner et al., 2016) as well as a description of the baseline task (Rabipour et al., 2020) are described elsewhere. Briefly, all participants had at least one parent or multiple siblings diagnosed with sporadic AD or a condition suggesting Alzheimer’s-like dementia within 15 years (Tschanz, Norton, Zandi, & Lyketsos, 2013). At baseline and during each subsequent follow-up assessment, participants performed neuropsychological tests as well as an object-location memory task in the scanner, described below. Here we focus on baseline analyses of the task-related fMRI based on self-identified biological sex. For more information on PREVENT-AD, see: douglas.qc.ca/page/prevent-alzheimer-the-centre.

### 2.3 Determination of Family History of AD

A brief questionnaire from the Cache County Study on Memory Health and Aging (Utah, USA) determined that all participants had a parent or multiple siblings: i) who had troubles with memory or concentration that was sufficiently severe to cause disability or loss of function; ii) for whom the condition had insidious onset or gradual progression and was not an obvious consequence of a stroke or other sudden insult.

### 2.4 APOE Genotyping

Genetic characterization was completed via blood draw, as previously described (Gosselin et al., 2016). DNA was isolated from 200 µl of the blood sample using QIASymphony and the DNA Blood Mini QIA kit (Qiagen, Valencia, CA, USA). *APOE* gene variant was determined using pyrosequencing with PyroMark Q96 (Qiagen, Toronto, ON, Canada). We defined -*APOE4* as *APOE* e3/3 genotype and +*APOE4* as *APOE* e3/4 genotype.

### 2.5 Neuropsychological Testing

Neuropsychological assessments took approximately 40 minutes to administer and were completed prior to every testing session. The test battery included:

*The Alzheimer-Dementia Eight Scale (AD8)*, an eight-item screening tool. The AD8 items index memory, orientation, judgment, and function. A score of two or above suggests impaired cognitive function (Galvin et al., 2005).

*The Clinical Dementia Rating (CDR)*, a five-point scale used to characterize memory, orientation, judgment & problem solving, community affairs, home & hobbies, and personal care (Berg, 1984). The information for each rating is obtained through a semi-structured interview of the patient and a reliable informant or collateral source (e.g., family member).

*The Montreal Cognitive Assessment (MoCA)*, a brief cognitive screening tool sensitive to mild declines in cognitive function (Nasreddine et al., 2004).

*The Repeatable Battery for the Assessment of Neuropsychological Status (RBANS)*, a battery of neuropsychological assessments aiming to identify abnormal cognitive decline in older adults (Randolph, Tierney, Mohr, & Chase, 1998). The RBANS provides scaled scores for five cognitive indices: immediate memory, visuospatial construct, language, attention, and delayed memory. We included these scaled scores, as well as the total score, in our analyses. Different versions of the RBANS were used in follow-up sessions to prevent practice effects.

### 2.6 Task fMRI: Behavioral Protocol

Participants were instructed to lie supine in a 3T Siemens Trio scanner (see below), while performing a source memory task programmed in E-Prime version 1.0 (Psychology Software Tools, Inc). During an initial encoding phase, participants were cued (10s) to memorize a series of 48 colored line drawings of common objects from the BOSS database (Brodeur, Guerard, & Bouras, 2014) in their spatial location (i.e., to the left or right of a central fixation cross). Each object was presented for 2000ms followed by a variable inter-trial interval (ITI; durations of 2200, 4400, or 8800ms; mean ITI = 5.13s) to add jitter to the fMRI data collection (Dale & Buckner, 1997). Following the encoding phase, there was a 20-min delay during which participants received structural MRI scans.

Following the 20-minute delay, a cue (10s) alerted participants to the beginning of the retrieval phase. During retrieval, participants were presented with 96 colored drawings of common objects: 48 ‘old’ (i.e., previously encoded) stimuli and 48 novel objects, in randomized order. Each object was presented in the center of the screen for 3000ms, with variable ITI (2200, 4400, or 8800ms). All participants used a fiber-optic 4-button response box to make task-related responses, and had an opportunity to familiarize themselves with the response choices during a practice session prior to testing. For each retrieval object, participants made a forced-choice between four-alternative answers: i) *“The object is FAMILIAR but you don’t remember the location”*; ii) *“You remember the object and it was previously on the LEFT”*; iii) *“You remember the object and it was previously on the RIGHT”*; and iv) *“The object is NEW”*. Thus (i) responses reflected object recognition, which may be more related to the utilization of familiarity vs. recollection based retrieval processes and reflected source failures, (ii) and (iii) responses reflect associative recollection of object-location associations (i.e., source hits) *if* the location endorsed was correct, or source misattributions (i.e., source failures) *if* the location endorsed was incorrect and (iv) responses reflected either correct rejections of novel objects or failed retrieval (“misses”). Responding (i)-(iii) to new objects reflected false alarms.

### 2.7 fMRI data acquisition

Functional magnetic resonance images were acquired with a 3T Siemens Trio scanner using the standard 12-channel head coil, located at the Douglas Institute Brain Imaging Centre in Montreal, Canada. T1-weighted anatomical images were acquired after the encoding phase of the fMRI task using a 3D gradient echo MPRAGE sequence (TR=2300 msec, TE=2.98 msec, flip angle=9°, 176 1mm sagittal slices, 1×1×1 mm voxels, FOV=256 mm). Blood Oxygenated Level Dependent (BOLD) images were acquired using a single-shot T2* -weighted gradient echo-planar imaging (EPI) pulse sequence with TR=2000 msec, TE=30 msec, FOV=256 mm. Brain volumes with 32 oblique slices of 4mm thickness (with no slice gap) were acquired along the anterior-posterior commissural plane with in-plane resolution of 4×4 mm.

A mixed rapid event-related design was employed to collect task-related blood oxygen level dependent (BOLD) activation during performance of the memory task (see above). Visual task stimuli were generated on a computer and back-projected onto a screen in the scanner bore. The screen was visible to participants lying in the scanner via a mirror mounted within the standard head coil. Participants requiring correlation for visual acuity wore plastic corrective glasses.

### 2.8 Data Analysis

#### 2.8.1 Preprocessing of fMRI data

We converted reconstructed images to NIfTI format and preprocessed them using in Statistical Parametric Mapping software version 12 (SPM12). Images from the first 10s of scanning were discarded to allow equilibration of the magnetic field. All functional images were realigned to the first image and corrected for movement artifacts using a 6-parameter rigid body spatial transform and a partial least squares approach. Functional images were then spatially normalized to the MNI EPI-template using the “Old Normalize” method in SPM12 at 4×4×4 mm voxel resolution, and smoothed using an 8 mm full-width half-maximum (FWHM) isotropic Gaussian kernel. Participants with head motion exceeding 4mm in the x, y, or z axis during encoding and retrieval were excluded from further analyses. Participants with movements that could not be sufficiently repaired, resulting in distorted brain images as judged by an examiner, were excluded from further analysis. To be included in further analyses, all participants were required to have a minimum of eight observations per event type (i.e., object recognition and source recollection).

#### 2.8.2 Behavioral analyses

We performed behavioral data analyses on neuropsychological tests and episodic memory task performance using JASP version 0.9.2, R version 3.4.1, and Psychometrica (available via: www.psychometrica.de). We used a significance threshold of *p*=0.05 with Greenhouse-Geisser corrections for sphericity and Holm-Bonferroni corrections for multiple comparisons, where applicable.

##### 2.8.2.1 Neuropsychological tests

We tested for sex differences in AD8, MoCA, and CDR scores using independent samples t-tests, and RBANS subscale scores using multivariate analysis of variance (MANOVA). We included self-reported sex (female, male) as the independent factor and test scores as the dependent variables.

##### 2.8.2.2 Episodic memory task

We calculated performance scores and mean reaction time (RT) for men and women, for the following response types: (i) correct object-only recognition with source failure (referred to as Source Failure) when participants correctly recognized old objects but could not recollect the spatial source (endorsed ‘familiar’) and/or when they correctly recognized an object but endorsed the wrong spatial source (source misattribution); (ii) correct object-location source retrieval, which we refer to as Source Hits; (iii) false alarms (incorrectly identifying new objects as old); (iv) misses (incorrectly identifying old objects as new); and (v) correct rejections (correctly identifying new objects). We computed Total Hits using the sum of Source Hits + Source Failures, and then calculated the proportion of Source hits/Total Hits and proportion of Source Failures/Total Hits, which reflected object-only retrieval. We further examined d’, computed as overall standardized hit rate minus standardized false alarm rate, as a measure of sensitivity, and c, computed by multiplying the average of the standardized hit and false alarm rates by -1, to measure response bias (Stanislaw & Todorov, 1999). We used MANOVA to evaluate sex differences on raw accuracy and response time (RT) measures, and as well as the proportion of total hits that were correct object-location source associations (i.e., source hits) vs. object-only recognitions (i.e., source failures).

#### 2.8.3 fMRI analyses

We performed spatio-temporal partial least squares (PLS) analyses using PLSGUI software (https://www.rotman-baycrest.on.ca/index.php?section=84) to identify whole-brain spatially and temporally distributed patterns of brain activity across experimental conditions and related to performance (McIntosh, Chau, & Protzner, 2004; see description of PLS advantages above). We conducted mean-centered task partial least squares (T-PLS) to identify group similarities and differences in event-related activity during successful object-location associative encoding and retrieval, and behavior partial least squares (B-PLS) to examine group similarities and differences in the correlations between event-related activity and the proportion of total hits that were correct object-location source associations vs. object-only recognitions. Details on PLS have been published elsewhere (Krishnan, Williams, McIntosh, & Abdi, 2011; McIntosh & Lobaugh, 2004).

As previously described (Rabipour et al., 2020), for both T-PLS and B-PLS we averaged the event-related data for each participant across the entire time series. PLS fMRI data were stacked into a between group data matrix wherein participants were nested within event/task condition; conditions were nested within group; and data for the group of Men were stacked above data for the group of Women. The conditions within group were stacked as follows: i) encoding objects in which participants subsequently remembered object-location source associations (ENC-Source Hit); ii) encoding objects in which participants subsequently remembered only the object identified, but failed to recall spatial source information or endorsed the wrong spatial source (ENC-Source Fail); iii) retrieval objects for which participants correctly recalled object-location source associations (RET-Source Hit); and iv) retrieval objects for which participants correctly recalled only the object identity, but failed to recall spatial source association or endorsed the wrong spatial source (RET-Source Fail). The stacked data matrix contained the fMRI data for each event onset (time lag = 0) with seven subsequent time lags, representing a total of 14s of activation after event onset (TR = 2s ⍰ 7 = 14 s). All participants analyzed had a minimum of eight correct events per event/condition type. There was no signal at lag 0 because data were baseline corrected to the event onset. Therefore, signal in subsequent lags was expressed as percentage deviation from event onset.

##### 2.8.3.1 Task PLS (T-PLS)

We mean centered the fMRI data column-wise during the encoding and retrieval phases of the episodic memory task, to evaluate whole-brain similarities and differences in brain activity related to encoding and retrieval of object-location associations in men compared to women. PLS performs singular value decomposition on the stacked data matrix to express the cross-covariance between the fMRI data and each condition into a set of mutually orthogonal latent variables (LVs). The number of LVs produced is equivalent to the number of event/task/condition types included in the analysis. Thus, this analysis yielded eight LVs (4 event-types * 2 groups). Each mean-centered LV comprises: i) a singular value reflecting the amount of covariance accounted for by the LV; ii) a design salience, depicted as a set of contrasts in the design salience plots (see results) that represent the relationship between tasks in each group and the pattern of brain activation; and iii) a singular image representing the numerical weights or “brain saliences” assigned to each voxel at each TR/time lag (i.e., the contribution of a region at each TR), yielding a spatio-temporal pattern of whole-brain activity that corresponds to the contrast effect identified by the design salience plot. Design saliences and brain saliences can be either positive or negative: positive brain saliences (depicted as warm-colored regions in the singular images) are positively correlated to *positive* design saliences, whereas negative brain saliences (depicted as cool-colored regions in the singular images) are positively correlated with *negative* design saliences (and vice-versa; Krishnan et al., 2011; McIntosh & Lobaugh, 2004). Thus, the pattern of whole brain activity shown in the singular image is symmetrically associated with the contrast effect identified by the design salience plot.

##### 2.8.3.2 Behavior PLS (B-PLS)

We used B-PLS to analyze whole-brain similarities and differences in brain activity directly correlated with source hits and source failures during encoding and retrieval between men and women. We stacked the behavioral vector containing source hits vs. failures as a proportion of total hits in the same order as the fMRI data matrix (i.e., participant within group). As in the T-PLS, B-PLS performed singular value decomposition of the stacked cross-correlation matrix to yield eight LVs. However, rather than design saliences, B-PLS analysis yields: i) a singular value, reflecting the amount of covariance explained by the LV; ii) a singular image consisting of positive and negative brain saliences, and iii) a correlation profile depicting how participants’ retrieval accuracy (proportion of source hits vs. failures) correlates with the pattern of brain activity identified in the singular image. The correlation profile and brain saliences represent a symmetrical pairing of brain-behavior correlation patterns for each group to a pattern of brain activity, respectively. As with the T-PLS analysis, brain saliences can have positive or negative values, and reflect whether activity in a given voxel is positively or negatively associated with the correlation profile depicted. Thus, negative correlations on the correlation plot indicate a negative correlation between performance and *positive* brain saliences (depicted as warm-colored regions in the singular image), but a *positive* correlation between performance and negative brain saliences (depicted as cool-colored regions in the singular image). Conversely, positive correlations indicate a positive correlation between performance and *positive* brain saliences, but a *negative* correlation between performance and negative brain saliences.

We assessed the significance of each LV in the T-PLS and B-PLS through 1000 permutations involving resampling without replacement from the data matrix to reassign the order of event types within participant. We determined the stability of the brain saliences using 500 bootstrap samples to calculate standard errors of voxel saliences for each LV, sampling participants with replacement while maintaining the order of event types for all participants. Significant voxels were those with bootstrap ratios ≥3.28 (positive salience brain regions) or ≤ - 3.28 (negative salience brain regions), corresponding to *p*<0.001, with a minimum spatial extent of 10 contiguous voxels.

We further computed temporal brain scores for each significant LV of the T-PLS and B-PLS. Similar to factor scores, temporal brain scores determine how strongly each participant’s data reflect the pattern of brain activity expressed in the singular image in relation to its paired design salience (T-PLS) or correlation profile (B-PLS), at each time lag (McIntosh, Chau, & Protzner, 2004). We report only peak coordinates from time lags at which the correlation profile maximally differentiated within the temporal window sampled (lags 2-5; 4-10s after event onset). We converted these peak coordinates to Talairach space using the icbm2tal transform (Lancaster et al., 2007) as implemented in GingerAle 2.3 (Eickhoff et al., 2009). Because our acquisition incompletely acquired the cerebellum, peak coordinates from this region are not reported. We used the Talairach and Tournoux atlas (Talairach & Tournoux, 1998) to identify the Brodmann area (BA) localizations of significant activations.

##### 2.8.3.3 Post hoc analyses

We further sought to investigate the potential influence of *APOE4* genotype on task-related brain activity and brain-behavior correlations in women compared to men. Because of our limited sample size, we performed only exploratory post hoc analyses of interactions between sex and *APOE4* carrier status (i.e., *-APOE4* vs. *+APOE4*) on brain scores identified in our T-PLS and B-PLS analyses, for encoding and retrieval of “old” objects. For T-PLS, we conducted repeated measures MANOVA to examine differences in brain scores within each significant LV, at each task condition (i.e., source recall vs. object recognition at encoding vs. retrieval phase), with sex and *APOE4* carrier status as between-subject factors. We followed up with repeated measures ANOVA and t-tests to examine significant effects revealed within each LV. For B-PLS, we explored differences in the correlations between brain scores and performance scores based on sex and *APOE4* carrier status, at each task condition. We used a significance threshold of *p*=0.05, and applied Greenhouse-Geisser corrections for sphericity and Bonferroni corrections for multiple comparisons, where applicable.

## 3. Results

Women and men were matched in demographics, including age and years of education. (Table 1a). In addition, we found no significant sex differences in body mass index (BMI), proximity to the age of parental AD onset (estimated years to AD onset; EYO), presence of AD-related biomarkers including the ratio of tau:Aβ in the cerebrospinal fluid, number of participants undergoing hormone replacement therapy, and motion during the fMRI scans (t_(77.5)_≤1.53, *p*≥0.13).

**Table 1.a).** Participant demographics represented as mean values ± standard deviation.

|  | **Women** | **Men** |
| --- | --- | --- |
| **Sample size (n) +APOE4** | 21 (53%) | 16 (40%) |
| **Age** | 63.13 ± 3.90 | 62.94 ± 3.74 |
| **BMI** | 25.92 ± 4.52 | 27.34 ± 3.79 |
| **Years of Education** | 15.40 ± 3.75 | 15.43 ± 3.06 |
| **Age proximity to parental diagnosis (EYO)** | 10.77 ± 7.44 | 9.31 ± 8.29 |
| **Tau:Aβ Ratio** | 0.26 ± 0.13 | 0.36 ± 0.27 |
| **Hormone Replacement Therapy (n)** | 7 (20%) | 2 (5%) |
| **Hysterectomy (n)** | 8 (20%) | - |
| **fMRI motion** | 0.044 ± 0.18 | 0.04 ± 0.10 |

*Welch's independent samples t-tests indicated no significant sex differences in mean demographic variables ( $t_{(77.5)} \leq 1.53$ , $p \geq .13$ ). Cerebrospinal fluid data not available in 22 women and 27 men.*

**Table 1.b).** Neuropsychological performance represented as mean values ± standard deviation.

|  | **Women** | **Men** |
| --- | --- | --- |
| **MoCA** | 28.50* ± 1.26 | 27.40 ± 1.80 |
| **AD8** | 0.08 ± 0.27 | 0.25 ± 0.63 |
| **CDR** | 0.00 ± 0.00 | 0.04 ± 0.13 |
| **Immediate Memory** | 104.54 ± 11.98 | 104.64 ± 10.06 |
| **Visuospat. Constr.** | 95.87 ± 13.25 | 91.97 ± 13.27 |
| **Language** | 100.13 ± 9.03 | 99.03 ± 8.07 |
| **Attention** | 106.85 ± 15.54 | 104.31 ± 14.62 |
| **Delayed Memory** | 101.97 ± 9.87 | 103.10 ± 9.20 |
| **RBANS Total** | 102.10 ± 10.59 | 100.39 ± 9.28 |

\* $t_{(78)} = 3.17$ , $p = .002$ , Cohen's $d = .71$
*MoCA = Montreal Cognitive Assessment; AD8 = Alzheimer-Dementia Eight Scale; CDR = Clinical Dementia Rating; RBANS = Repeatable Battery for the Assessment of Neuropsychological Status. RBANS data missing from one man and one woman who did not receive the test.*

**Table 1.c).** Episodic memory task performance and RT represented as mean values ± standard deviation.

|  | **Women** | **Men** |
| --- | --- | --- |
| **% Total Hits** | 85.21 ± 10.75 | 82.29 ± 12.72 |
| **% Source Hits** | 55.37 ± 13.69 | 52.14 ± 16.16 |
| **% Source Failures** | 17.66 ± 9.83 | 16.35 ± 12.29 |
| **% Source Misattributions** | 12.19 ± 6.27 | 13.80 ± 5.89 |
| **% Misses** | 14.79 ± 10.75 | 17.71 ± 12.72 |
| **% False Alarms** | 13.97 ± 10.07 | 15.68 ± 12.79 |
| **% Correct Rejections** | 86.25 ± 10.14 | 84.06 ± 12.82 |
| **Proportion Source Hits** | 0.64 ± 0.12 | 0.62 ± 0.14 |
| **Proportion Source Fails** | 0.36 ± 0.12 | 0.38 ± 0.14 |
| **d'** | 0.17 ± 1.38 | -0.26 ± 1.50 |
| **c** | -0.04 ± 0.59 | -0.002 ± 0.83 |
| **Source Hits (sec)** | 1.67 ± 0.31 | 1.79 ± 0.30 |
| **Source Failures (sec)** | 2.28 ± 0.61 | 2.45 ± 0.64 |
| **Source Misattributions (sec)** | 2.03 ± 0.50 | 2.17 ± 0.42 |
| **Misses (sec)** | 1.94 ± 0.48 | 2.13 ± 0.52 |
| **False Alarms (sec)** | 2.53 ± 0.55 | 2.45 ± 0.47 |
| **Correct Rejections (sec)** | 1.62 ± 0.28 | 1.68 ± 0.34 |

### 3.1 Neuropsychological Performance

Table 1b shows participant scores on the neuropsychological tests. We found a significant main effect of sex on MoCA total score, whereby women performed better than men (t_(78)_=3.17, *p*=0.002, Cohen’s *d*=0.71). Following up on this effect revealed that six men – but no women – scored below the clinical cutoff of 26 (*X*^2^=6.49, *p*=0.01). We found no significant effect of sex on AD8 or CDR scores (*p*>0.05). Similarly, MANOVA on RBANS subscales revealed no significant sex differences in RBANS performance (Wilk’s λ=0.96, F_(6,71)_=0.54, *p*=0.77).

### 3.2 fMRI: Episodic Memory Task Performance

The behavioral results from the object-location associative memory task are presented in Table 1c. The MANOVA revealed no significant sex differences in task performance (Wilk’s λ=0.94, F_(5,74)_=0.95, *p*=0.45) or RT (Wilk’s λ=0.89, F_(6,71)_=1.80, *p*=0.11). Between-groups (i.e., sex) repeated measures ANOVA examining differences in RT by response type revealed significant response type differences in RT (F_(3.94,295.3)_=67.73, *p*<0.0001, η_p_^2^=.48;). Both men and women had significantly longer RT for FA trials compared to source hits, source misattributions, misses, and correct rejections (t_(79)_ ≥6.95, *p*≤0.0001, Cohen’s *d*≥0.79), and significantly faster RT for correct rejections compared to all other trials (t_(79)_ ≥7.27, *p*≤0.0001, Cohen’s *d*≥0.83), with the exception of correct source hits. Of trials presenting “old” (i.e., previously viewed) objects, participants had the fastest RT for source hits (t_(79)_ ≥5.73, *p*≤0.0001, Cohen’s *d*≥0.65). Thus, source hits and correct rejection decisions were significantly faster than other decision types.

### 3.3 fMRI results

#### 3.3.1 T-PLS

The PLS analysis yielded four significant LVs (*p*≤0.04). The first significant LV (LV1, *p*<0.001) accounted for 48.16% of the cross-block covariance and identified brain regions in which activity significantly differed during correct rejections and encoding, compared to retrieval, in both groups (Figure 2A). Table 2 lists the local maxima from LV1. In both men and women, the positive salience brain regions were more active during retrieval, compared to encoding and correct rejections, whereas the negative salience brain regions were more active during encoding and correct rejections, compared to retrieval. Positive salience regions included bilateral medial and lateral prefrontal cortex (PFC), posterior cingulate and precuneus, inferior parietal, medial temporal, and lateral occipital cortices. Negative salience regions included bilateral ventromedial/orbital and superior PFC and lateral middle temporal cortices, and right fusiform cortex. Thus, this LV identified brain regions that were differentially activated to during the perception of novel objects (correct rejections) and the successful encoding of objects and object-location associations (source failures and hits, respectively), compared to the successful retrieval of previously seen objects and object-location associations. Consistent with this interpretation, post hoc analysis of LV1 confirmed a significant task main effect (F_(3.2,239.4)_=65.64, *p*<0.001, η_p_^2^=0.46), and no other significant effects (Figure 3a).

**Table 2.**
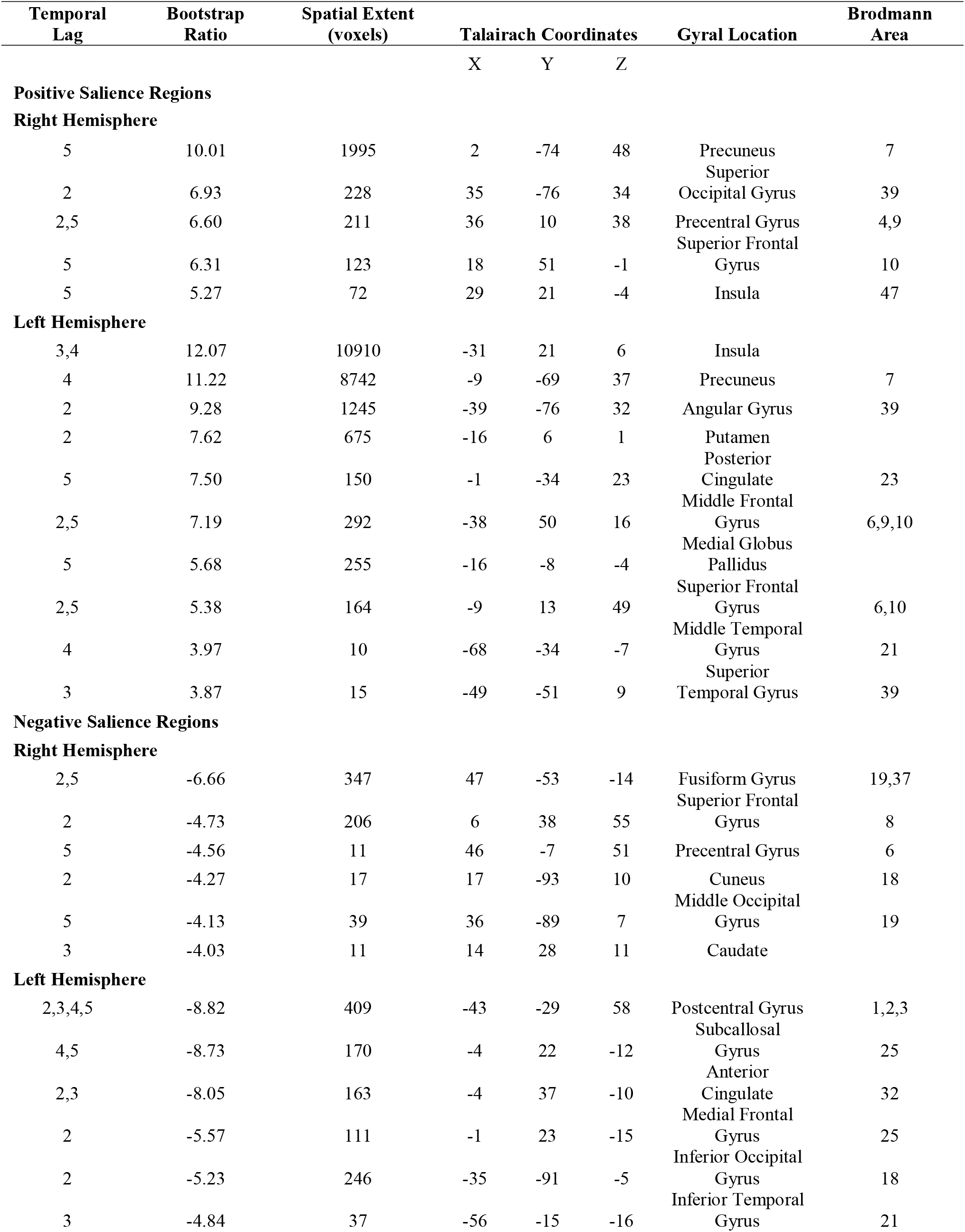
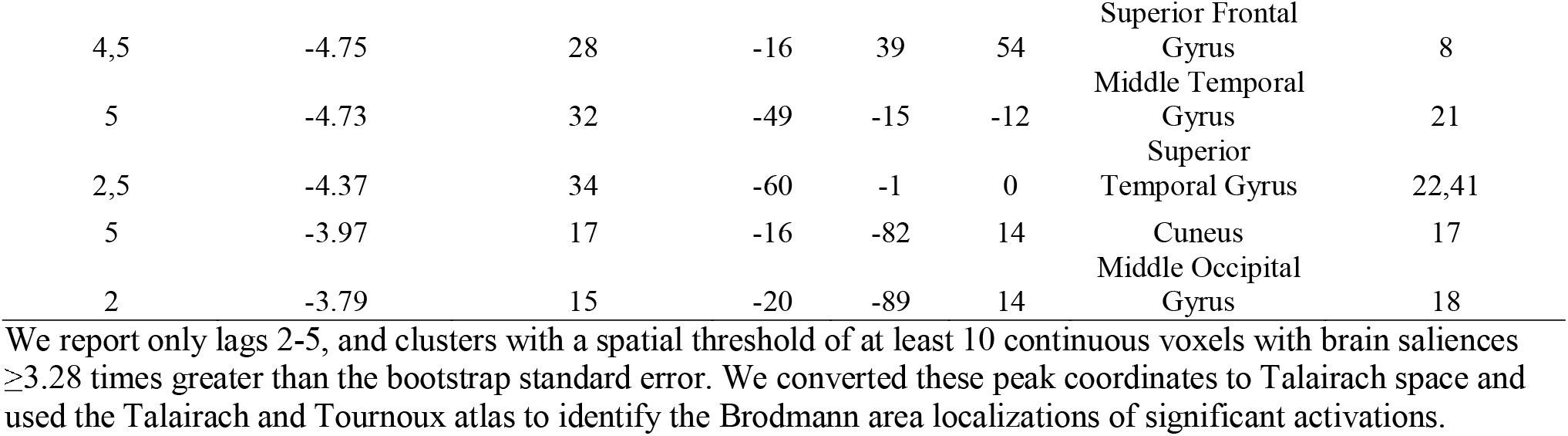
Local maxima revealed for LV1 of the T-PLS analysis.

**Fig. 1.**
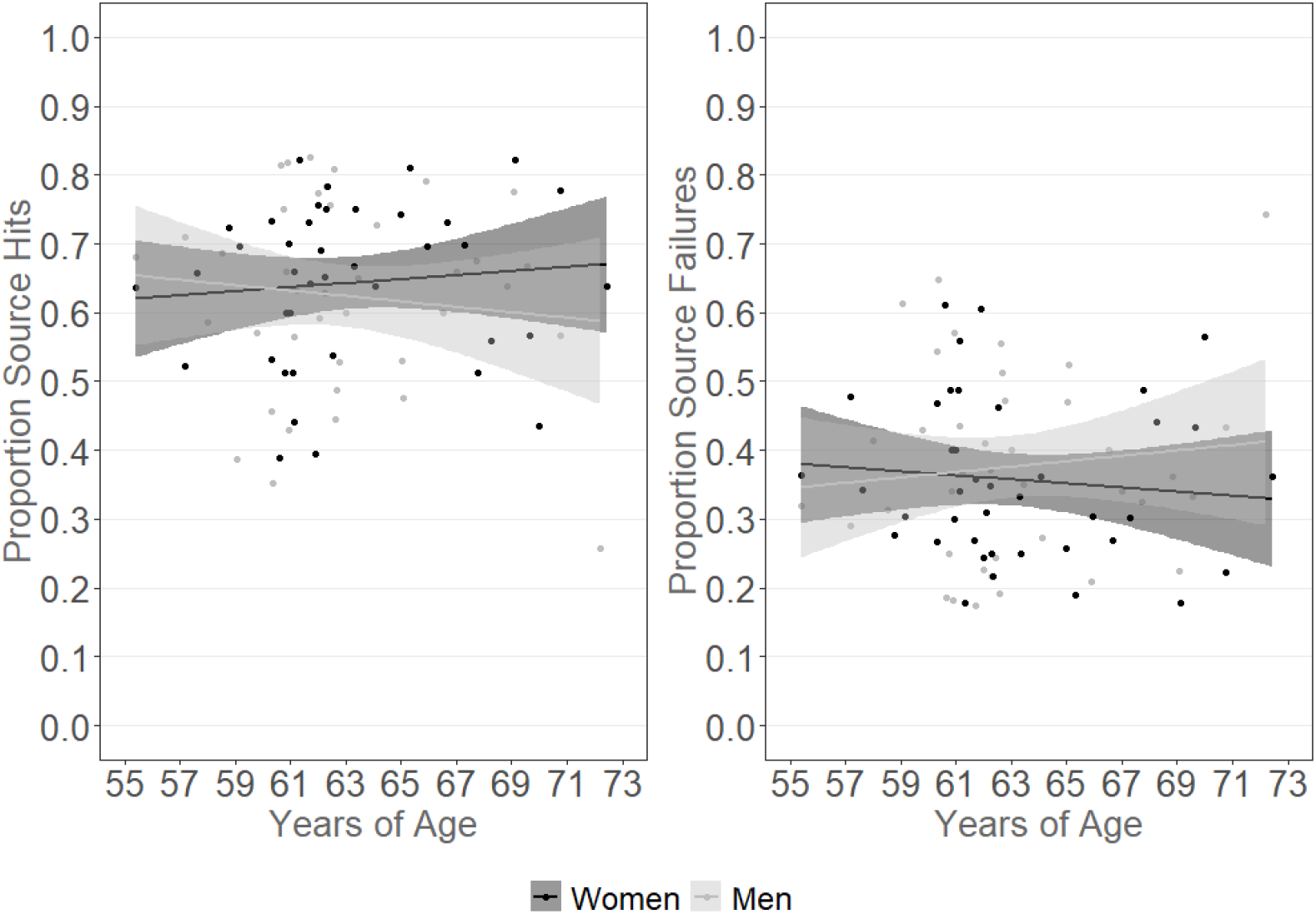
Source hits and source failures, calculated as a proportion of total hits, in women and men. Shaded regions represent 95% CI.

**Fig. 2.**
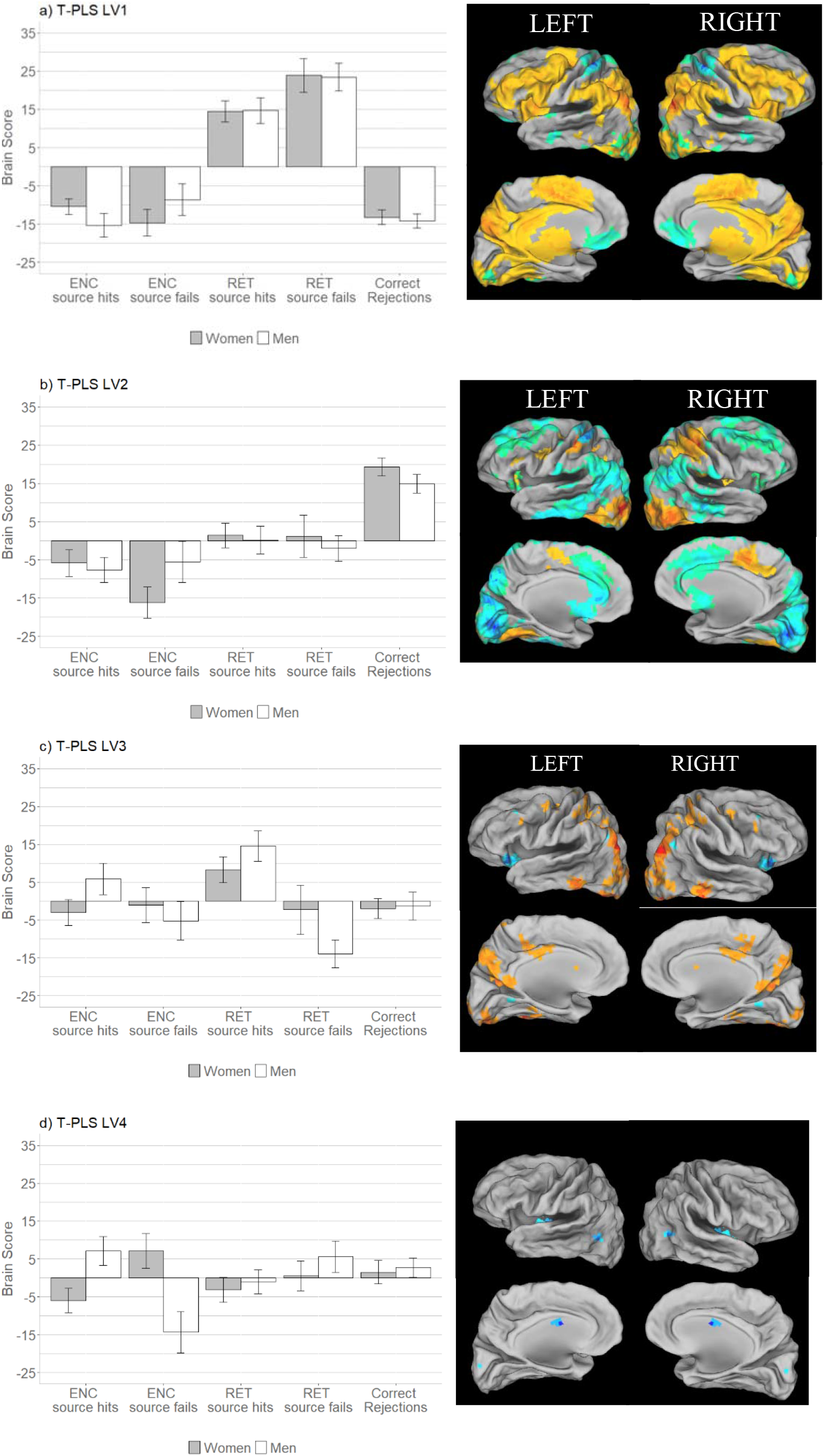
Design salience plot and singular image representing brain activity patterns by condition for (a) LV1, (b) LV2, (c) LV3, and (d) LV4 revealed by the T-PLS analysis. LEFT: Error bars on design salience plots represent standard error of the mean. Positive brain scores indicate conditions in which activity was greater in positive brain salience regions (shown in warm tones in the singular images) and vice versa. Negative brain scores indicate conditions in which activity was greater in negative brain salience regions (shown in cool tones in the singular images) and vice versa. RIGHT: Singular images representing peak coordinates thresholded at a bootrstrap ratio of ±3.28. Warm-toned brain regions represent positive brain saliences; cool-toned regions represent negative brain saliences. Activations are presented on template images of the lateral and medial surfaces of the left and right hemispheres of the brain using Caret software (http://brainvis.wustl.edu/wiki/index.php/Caret:Download).

**Fig. 3.**
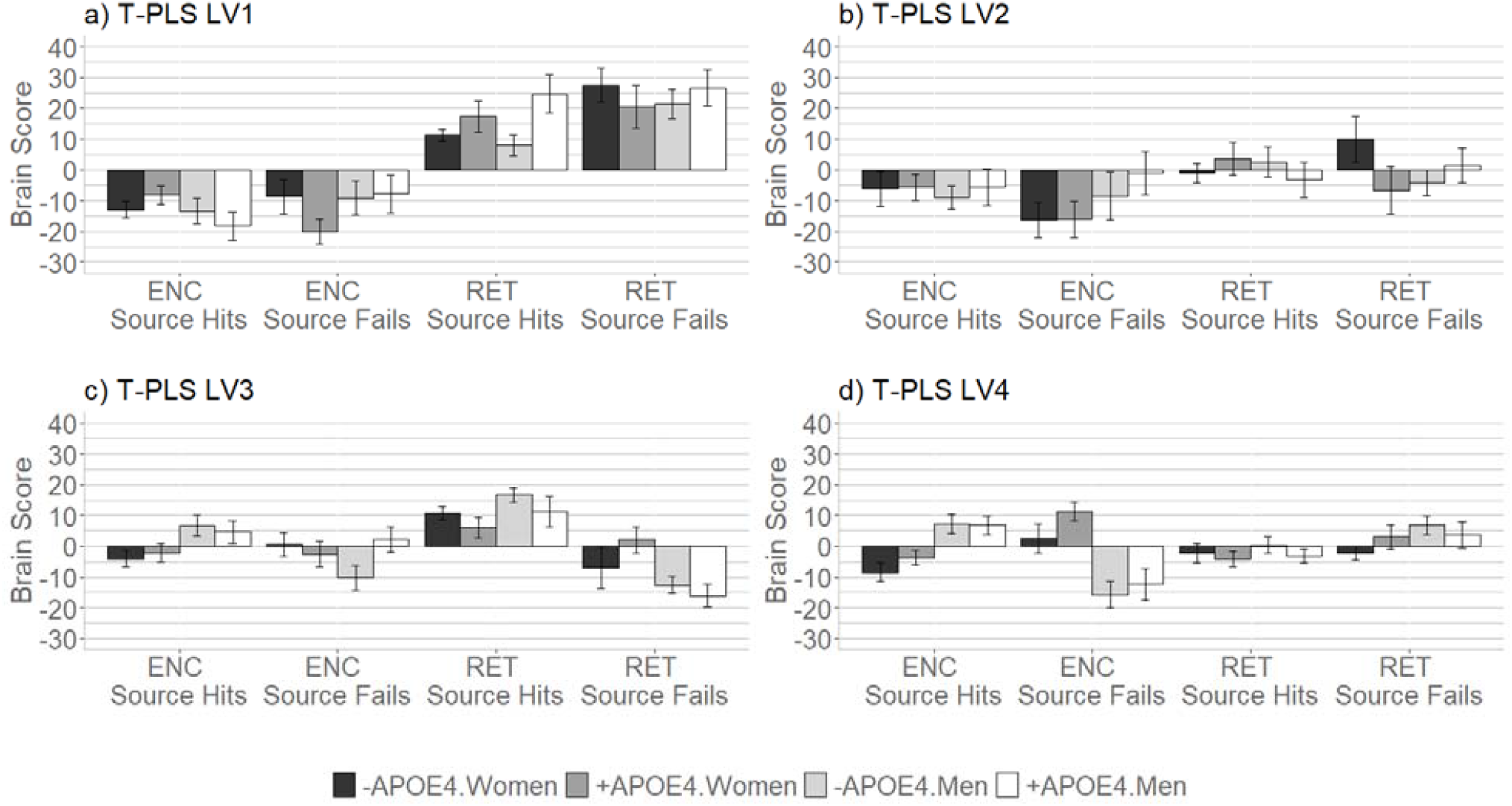
Design salience plots depicting brain scores by sex and *APOE4* for (a) LV1, (b) LV2, (c) LV3, and (d) LV4 of the T-PLS analysis, corresponding to the post hoc analyses. Error bars represent standard error of the mean.

**Fig. 4.**
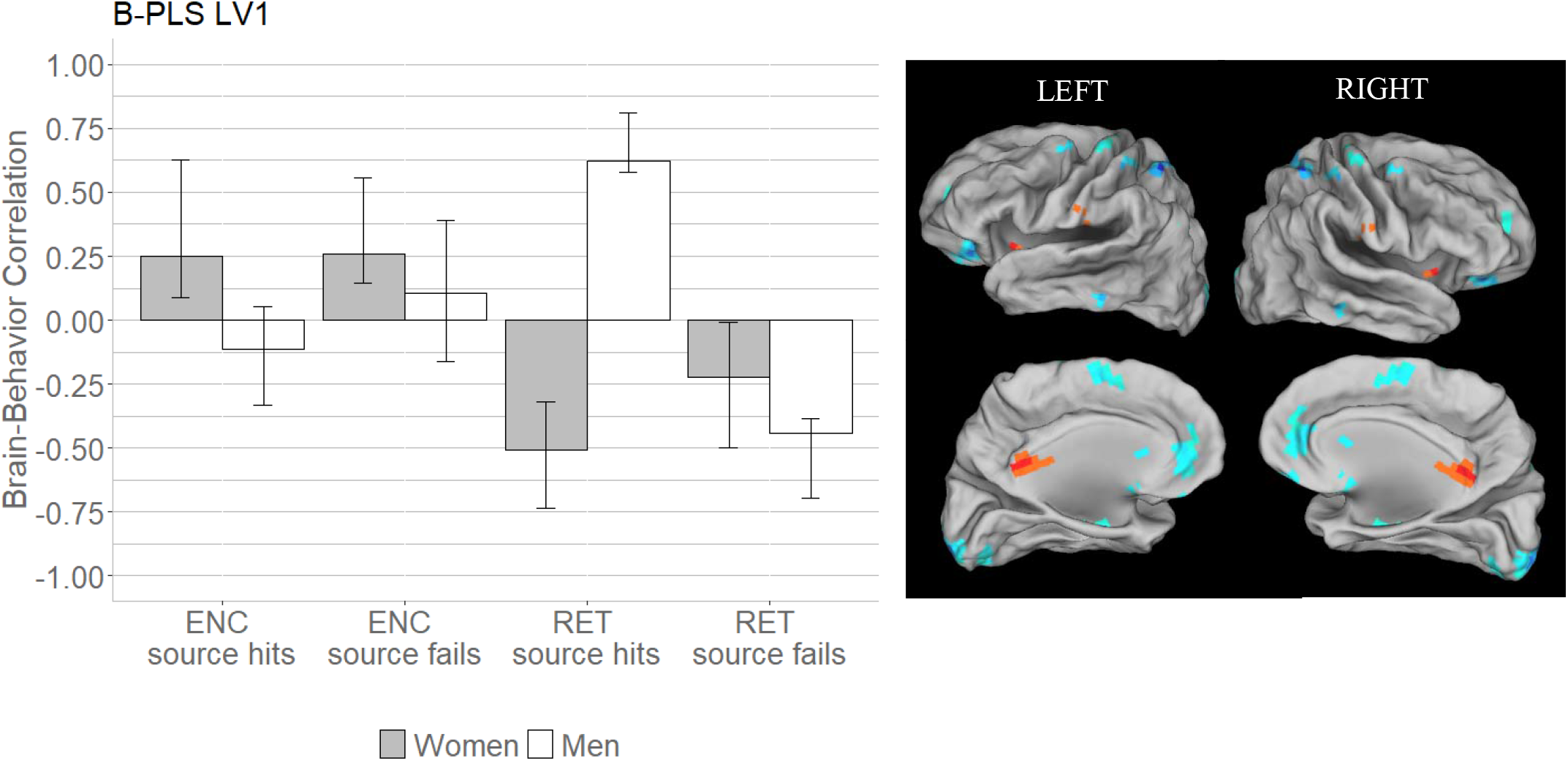
Correlations between brain activity and task performance by condition, revealed by the only significant LV of the B-PLS analysis. LEFT: Bars represent brain-behavior correlations for each group, by condition. Positive correlations indicate conditions in which performance was positively associated with activity in positive brain salience regions (shown in warm tones in the singular images) and vice-versa. Negative correlations indicate conditions in which performance was positively associated with activity in negative brain salience regions (shown in cool tones in the singular images) and vice-versa. Error bars represent standard error of the mean. RIGHT: The singular image thresholded at a bootstrap ratio of ±3.28, depicting the identified negative brain saliences in cool tones. Activations are presented on template images of the lateral and medial surfaces of the left and right hemispheres of the brain using Caret software (http://brainvis.wustl.edu/wiki/index.php/Caret:Download).

LV2 of the T-PLS accounted for 18.64% of the cross-block covariance (*p*<0.001). We present the local maxima from this LV in Table 3. The design salience plot and singular image presented in Figure 2B indicate that this LV identified brain regions differentially activated during the successful encoding of object-only information with source failure and successful encoding of object-location source information (negative salience brain regions), compared to correct rejections of new items (i.e., novelty detection; positive salience brain regions), in both sexes. Consistent with our interpretation, the post-hoc analysis identified only a significant task main effect (F_(3.6,272)_=15.79, *p*<0.001, ηp^2^=0.17; Figure 2b). Positive salience regions included bilateral precentral gyrus, occipital gyrus and right ventrolateral PFC. Negative salience brain regions included bilateral medial, orbital and dorsal/superior PFC, middle temporal, temporo-parietal, and medial occipital cortices, and the right parahippocampal cortex.

**Table 3.**
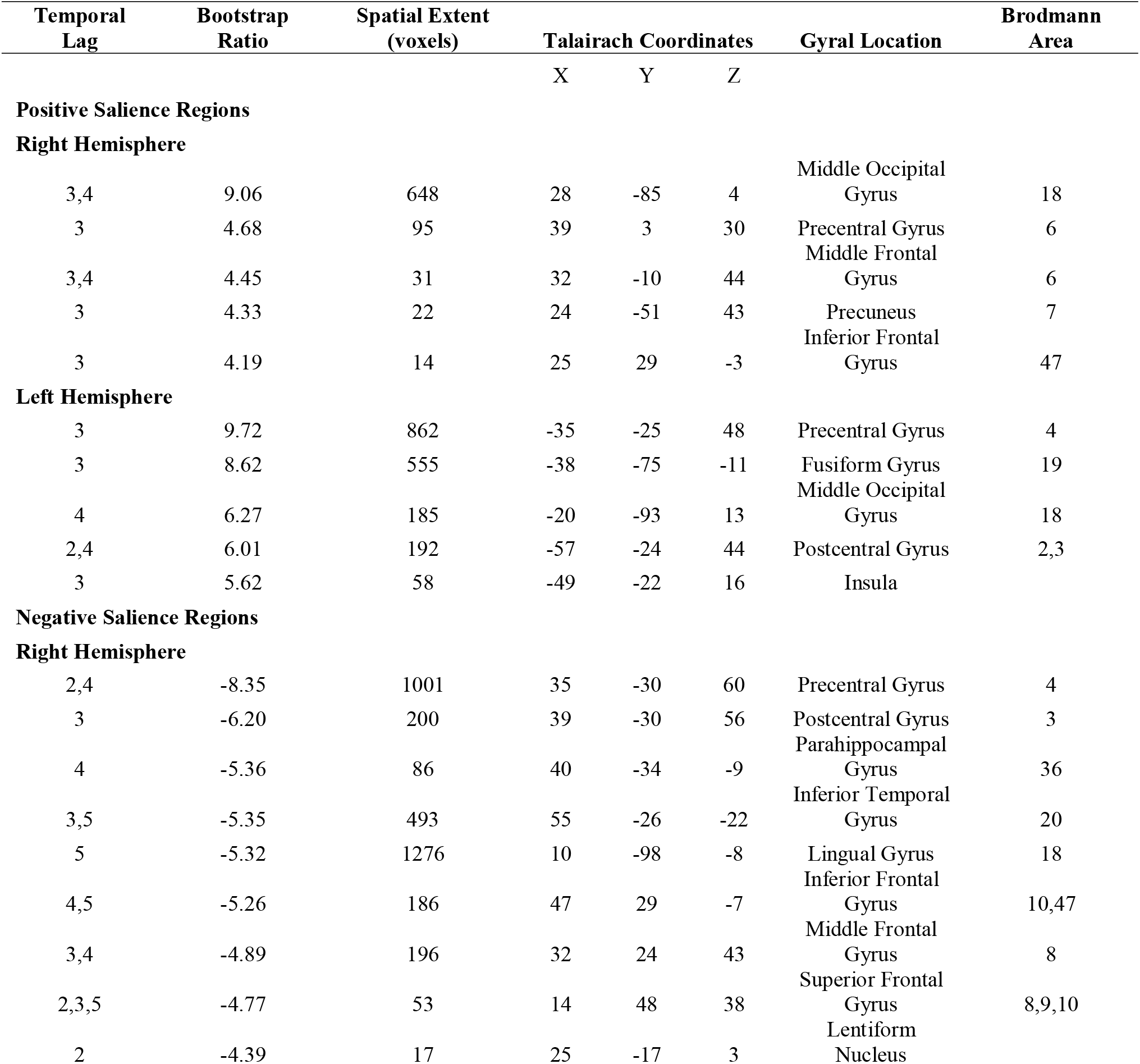
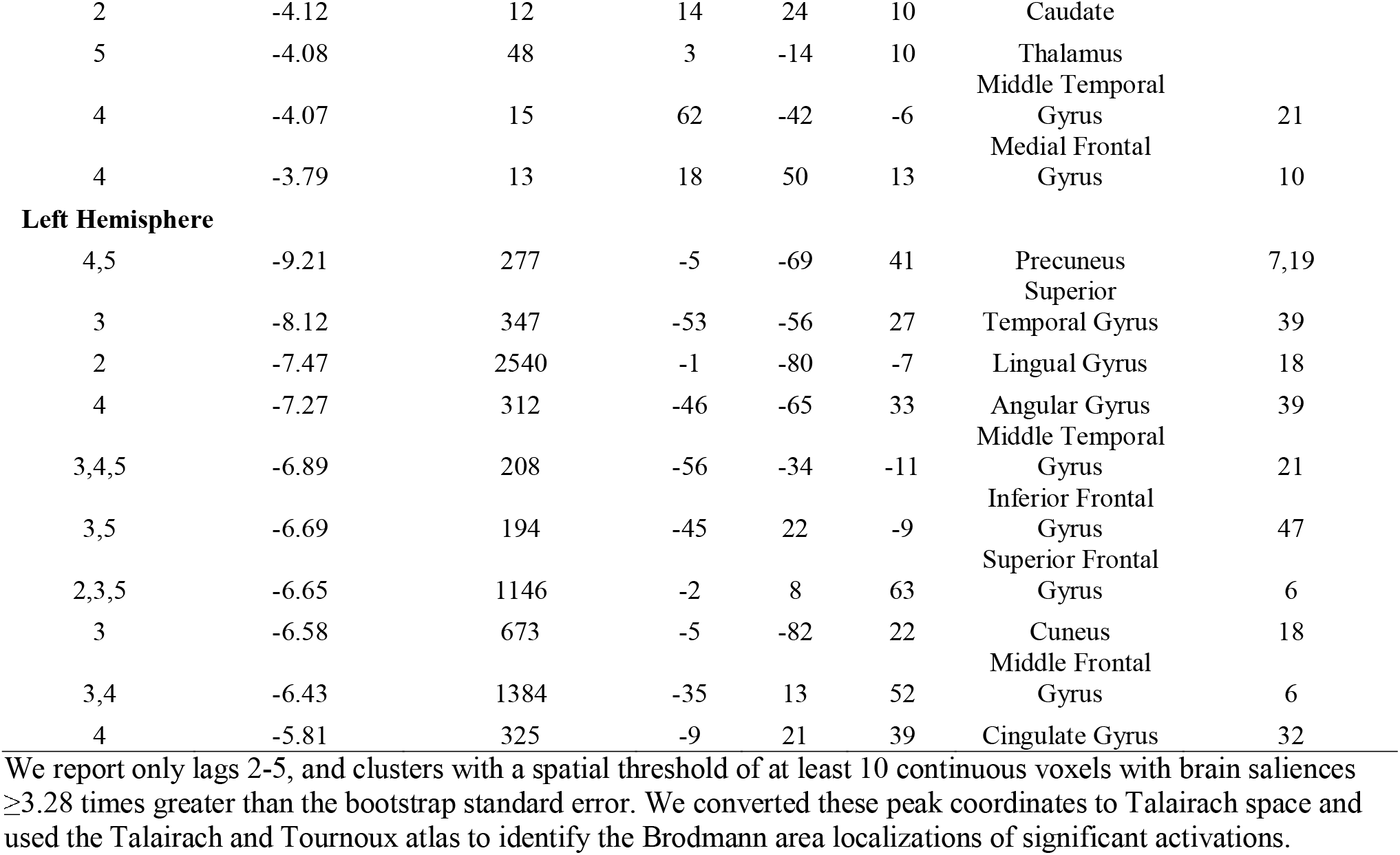
Local maxima revealed for LV2 of the T-PLS analysis.

LV3 of the T-PLS accounted for 10.52% of the cross-block covariance (*p*=0.004). The local maxima from this LV are presented in Table 4. The design salience plot indicates that LV3 identified sex differences in event-related activity during encoding and retrieval (Figure 2C). Specifically, positive salience brain regions were more active during the encoding and retrieval of object-location associative source information compared to the encoding and retrieval of object-only retrieval with source failure (negative salience brain regions) in men. Women exhibited the same pattern of increased activity in positive salience brain regions only during object-location associative source retrieval. Thus, this LV identified activations related to encoding and object-only retrieval that were specific to men. Positive salience brain regions included caudate, bilateral superior and middle temporal cortex, dorsal occipital cortex, medial cingulate, inferior parietal cortex, and precuneus. The few negative salience brain regions included bilateral ventrolateral PFC and insula. The post hoc analysis of LV3 confirmed there was a significant sex-by-event-type interaction (F_(3.6,273.6)_=5.87, *p*<0.001, η_p_^2^=0.07) as well as a significant three-way interaction between sex, *APOE4*, and event type (F_(3.6,273.6)_=2.81, *p*=0.03, η_p_^2^=0.04; Figure 3c). Specifically, compared to women, men demonstrated greater activation of positive salience brain regions during encoding of source hits (t_(78)_=2.81, *p*=0.006, Cohen’s *d*=0.63) and of negative salience regions during retrieval of source failures (t_(61.3)_=2.64, *p*=0.01, Cohen’s *d*=0.59). This latter effect appeared to be driven by *-APOE4* individuals, where *-APOE4* men demonstrated significantly greater activation of negative salience regions during retrieval of object-only information (i.e., source failures) compared to *-APOE4* women (t_(35)_=3.14, *p*=0.003, Cohen’s *d*=1.04).

**Table 4.**
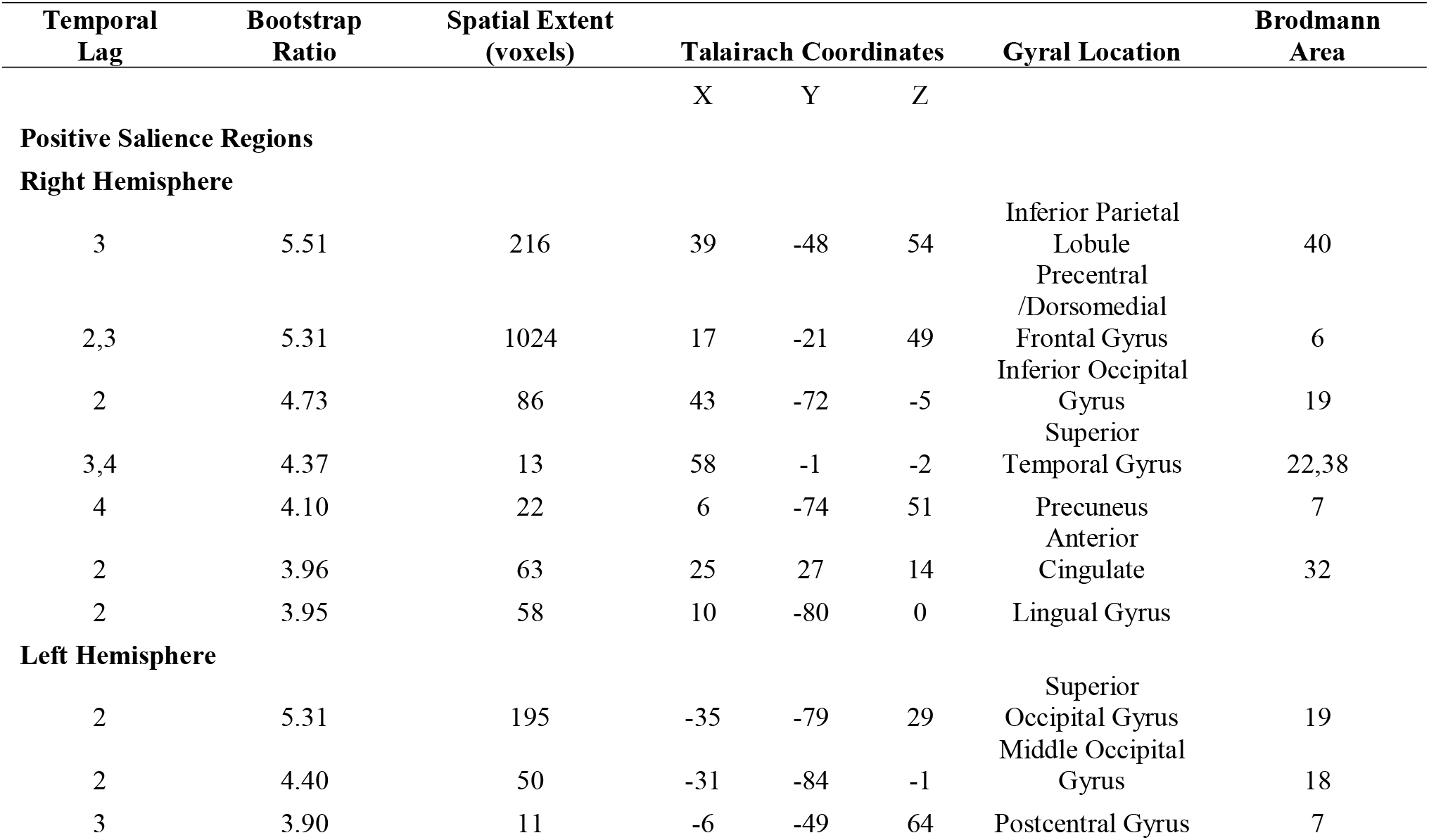
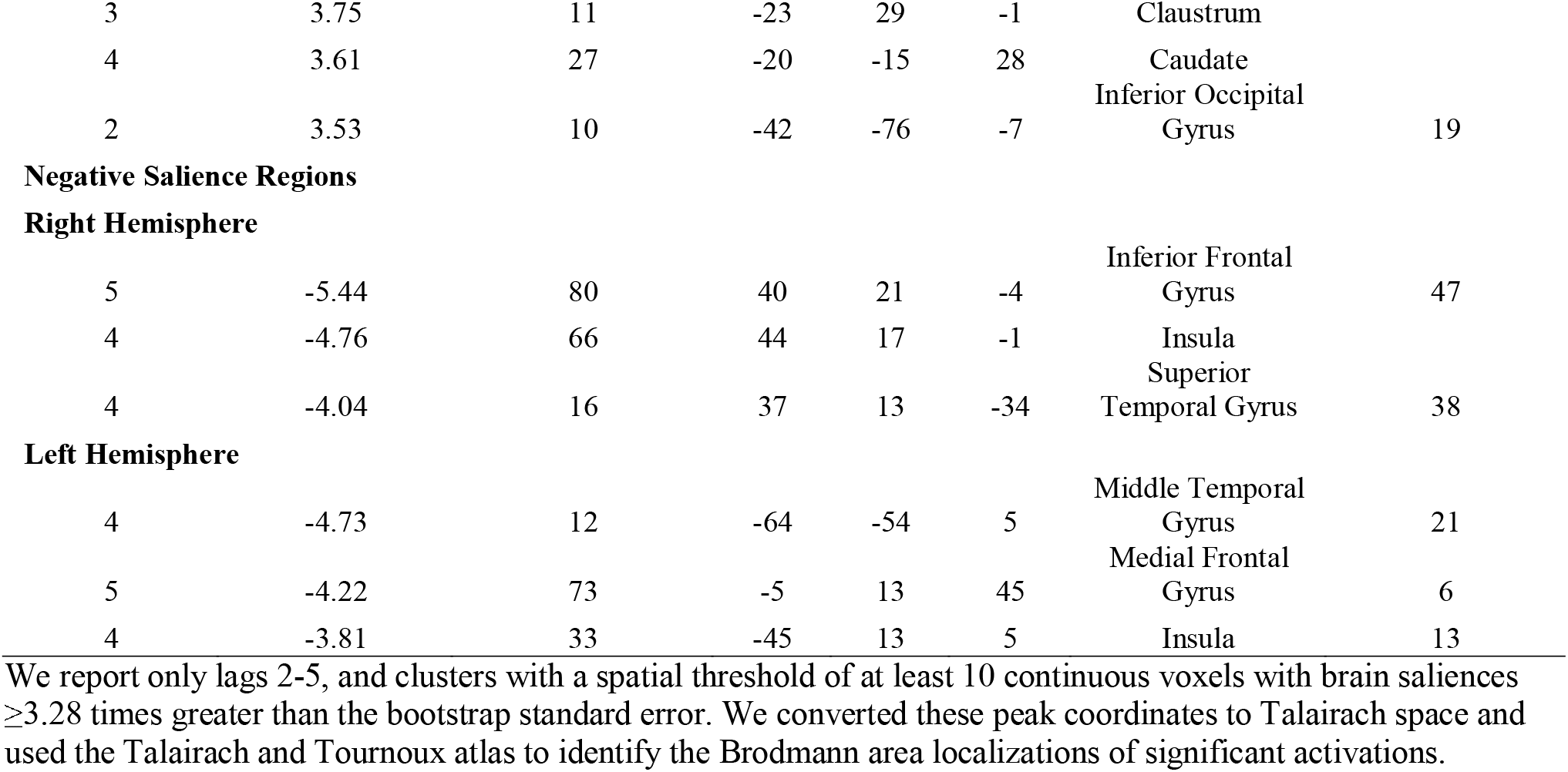
Local maxima revealed for LV3 of the T-PLS analysis.

LV4 of the T-PLS accounted for 7.49% of the cross-block covariance (*p*=0.04). The local maxima from this LV, all of which were negative, are presented in Table 5. The design salience plot indicates that LV4 identified sex differences in event-related activity during the encoding of object-location associative source hits vs. object-only encoding with source failure (Figure 2D). Specifically, the negative salience brain regions were more active during the encoding of subsequent source hits (i.e., object-location associations), compared to object-only encoding with source encoding failure in women. We found the opposite pattern in men, where negative salience brain regions were more active during object-only encoding (with failure of source encoding) compared to encoding of subsequent source hits and retrieval of object-only information. The negative salience brain regions included the bilateral cingulate, caudate, and left middle occipital cortex. Thus, this LV revealed distinct activations related to the encoding of source hits in women vs. source failures in men. Post hoc analysis of this LV (Figure 3d) supported this interpretation, indicating a significant sex-by-event-type interaction (F_(3.2,246.8)_=17.22, *p*<0.001, η_p_^2^=0.19). Specifically, women demonstrated significantly higher activation of negative salience brain regions during encoding of subsequent source hits (t_(78)_=4.08, *p*=0.002, Cohen’s *d*=0.46), and men demonstrated significantly higher activation of negative salience brain regions during encoding of subsequent source failures (t_(78)_=6.48, *p*<0.001, Cohen’s *d*=0.73). Because this LV accounted for so little variance and identified few significant activations, we do not discuss these results further.

**Table 5.** Local maxima revealed for LV4 of the T-PLS analysis.

| Temporal Lag | Bootstrap Ratio | Spatial Extent (voxels) | Talairach Coordinates |  |  | Gyrat Location | Brodmann Area |
| --- | --- | --- | --- | --- | --- | --- | --- |
|  |  |  | X | Y | Z |  |  |
| Negative Salience Regions |  |  |  |  |  |  |  |
| Right Hemisphere |  |  |  |  |  |  |  |
| 4 | -4.59 | 14 | 17 | 11 | 31 | Cingulate Gyrus | 24 |
| 2 | -4.37 | 83 | 29 | -17 | 7 | Putamen |  |
| Left Hemisphere |  |  |  |  |  |  |  |
| 2 | -4.31 | 16 | -42 | -66 | 5 | Middle Occipital Gyrus | 37 |
| 2 | -4.10 | 60 | -34 | -13 | 2 | Clastrum |  |
| 5 | -3.75 | 11 | -5 | 0 | 22 | Cingulate Gyrus | 24 |
| 5 | -3.62 | 11 | -20 | -37 | 22 | Caudate |  |

We report only lags 2-5, and clusters with a spatial threshold of at least 10 continuous voxels with brain saliences $\geq 3.28$ times greater than the bootstrap standard error. We converted these peak coordinates to Talairach space and used the Talairach and Tournoux atlas to identify the Brodmann area localizations of significant activations.

#### 3.3.2 B-PLS Results

The B-PLS yielded one significant LV (*p*=0.02), which accounted for 21.56% of the cross-block covariance and identified sex differences in brain-behavior correlations (Figure 3). Table 6 lists the local maxima for the significant brain regions. The brain-behavior correlation plot indicates that, in women, activity in positive salience brain regions during encoding correlated with better subsequent retrieval of both object-location associations and object-only information. In contrast, activity in negative salience brain regions during retrieval correlated with better retrieval of both source and object-only information. Thus, in women, this LV differentiated brain regions in which activity predicted successful encoding, compared to successful retrieval. In men, this LV identified brain regions in which activity during retrieval was differentially correlated with the retrieval of source information, compared to the retrieval of object-only information with source failures. Specifically, in men, activity in positive salience brain regions correlated with better source retrieval and activity in negative salience brain regions correlated with better object-only retrieval. Positive salience brain regions included bilateral insula/ventrolateral PFC, posterior cingulate, and left supramarginal gyrus. Negative salience brain regions included bilateral parahippocampal gyrus, anterior cingulate, precuneus and left medial occipital and ventrolateral PFC.

**Table 6.**
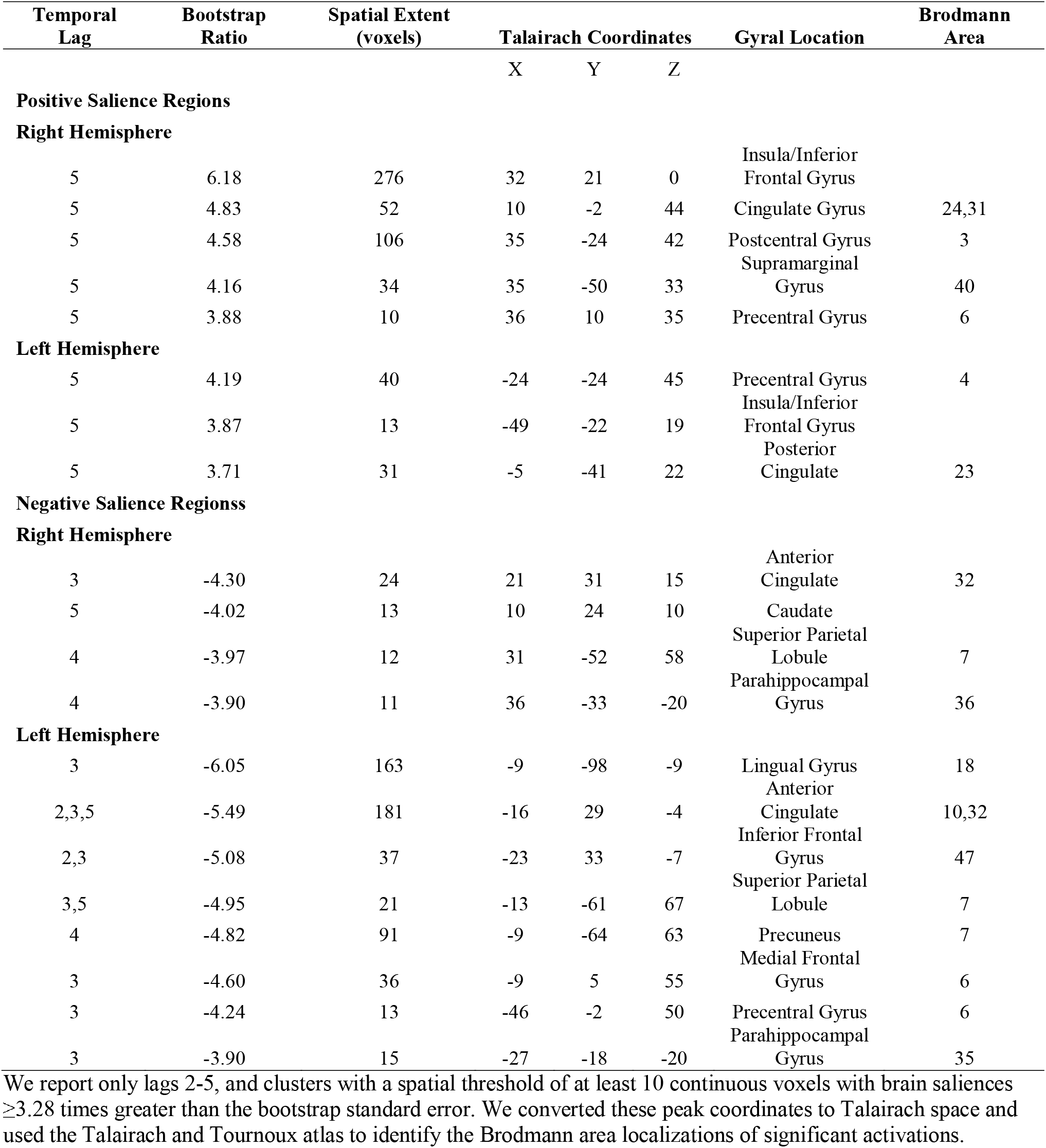
Local maxima revealed by LV1 of the B-PLS analysis.

The post hoc analysis of the B-PLS indicated a significant sex-by-event-type interaction during retrieval of object-location associations (Figure 5). Specifically, women demonstrated a negative correlation between performance and positive brain salience regions (r=-0.51), whereas men showed a positive correlation between performance and positive brain salience regions (r=0.62; Z=5.54, *p*<0.001).

**Fig. 5.**
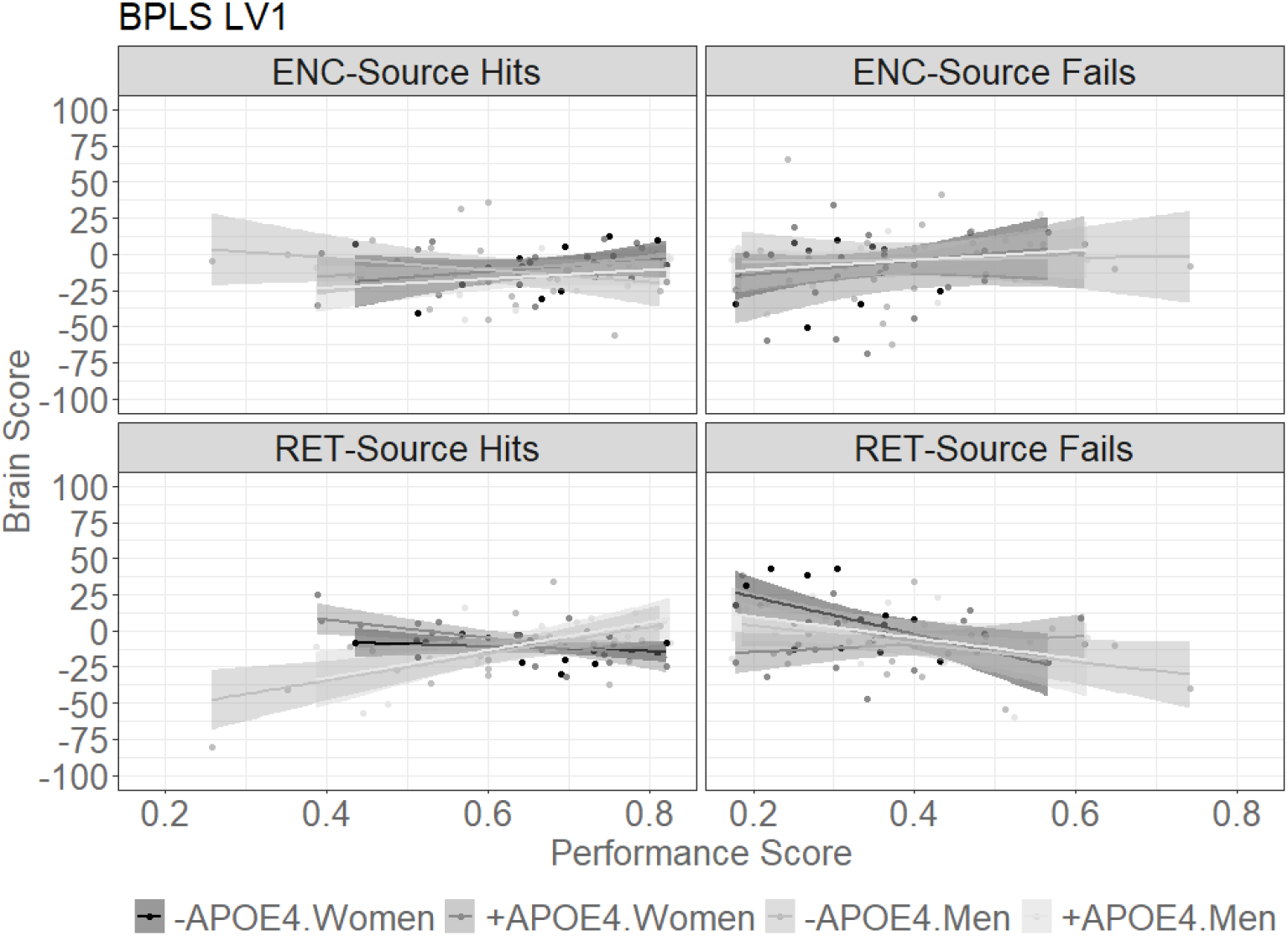
Sex by *APOE4* correlations between brain activity and task performance by condition, revealed by the B-PLS analysis. Shaded regions represent 95% CI.

## 4. Discussion

Evidence suggests that AD risk and AD-related declines in episodic memory and general cognition are higher in women compared to men (Gamache et al., 2020; Irvine et al., 2012; Mazure & Swendsen, 2016). Understanding the nature of the disproportionate impact of AD – on women, compared to men, therefore remains a priority (Alzheimer’s Association, 2020; Nebel et al., 2018). Drawing on baseline data from the PREVENT-AD dataset (Breitner et al., 2016), here we examined sex differences in the neural correlates of episodic memory performance in a sample of older adults who all had first-degree family history of AD. Building on our previous work (Rabipour et al., 2020), we assessed the relationship between self-reported biological sex and brain activity from the whole-brain perspective using PLS, a data-driven multivariate approach, and a novel episodic memory task distinguishing object-location source association from recognition. To account for the limited sample of men compared to women, we analyzed a partial sample wherein women were matched to men with respect to age, years of education, BMI, and estimated years to symptom onset.

### 4.1 Few Sex Differences in Behavior and Brain Activity Related to Encoding and Retrieval of Old vs. Novel Objects

Our analyses identified few sex differences in behavioral performance. Both women and men performed generally well on neuropsychological tests and in line with previously reported trends in this population (Larouche et al., 2016). We found no significant sex differences in performance on the episodic memory task, including in the accuracy and RT associated with object-location associations and object-only recognition. This comparability of behavioral performance between women and men is unsurprising given our prior findings in the larger PREVENT-AD baseline analysis (Rabipour et al., 2020).

The data-driven mean-centered T-PLS analysis identified general patterns of object-only and object-location associative encoding and retrieval activity that were common to both sexes (LV1, LV2) and supported the task-related brain activation patterns we previously found in this cohort (Rabipour et al., 2020). Women and men exhibited similar patterns of event-related brain activity during successful encoding vs. retrieval of object-only and associative information (LV1). Both groups demonstrated greater activity in bilateral ventromedial/orbital and superior PFC as well as lateral middle temporal cortices and right fusiform cortex during encoding, compared to broad bilateral activity in medial and lateral PFC, posterior cingulate and precuneus, inferior parietal, medial temporal, and lateral occipital cortices at retrieval. In addition, brain activation patterns were consistent across both sexes for the encoding of old objects compared to novelty detection (LV2). Women and men exhibited more activity in bilateral medial, orbital, and dorsal/superior PFC, middle temporal, temporo-parietal, medial occipital regions, and right parahippocampal cortex during the encoding of object-location associations and object-only information. Conversely, novelty detection associated with more activity in bilateral precentral and occipital regions and right ventrolateral PFC in both sexes.

The episodic memory-related brain activity patterns identified in T-PLS LV1 and LV2 are consistent with those previously reported for the encoding and retrieval of associative information in older adults (Maillet & Rajah, 2014; Salami, Eriksson, & Nyberg, 2012). In particular, our results support greater engagement of medial orbitofrontal and dorsal/superior PFC, lateral middle temporal, as well as primary and secondary sensory cortices (e.g., prefrontal and ventral occipito-temporal areas) and parahippocampal cortex during visual associative encoding (de Chastelaine, Mattson, Wang, Donley, & Rugg, 2015, 2016; Dennis et al., 2019; T. Sommer, Rose, Weiller, & Buchel, 2005) and of dorsal precuneus during visuospatial associative encoding (Rami et al., 2012; Schott et al., 2019). In comparison, during episodic retrieval we found activation of medial temporal, fronto-parietal control, and default-mode network regions such as the posterior cingulate and inferior parietal cortices (Huo, Li, Wang, Zheng, & Li, 2018; Sestieri, Corbetta, Romani, & Shulman, 2011). These patterns are consistent with the Retrieval, Experience, and Decision (RED) model (Kim, 2020), suggesting multi-stage engagement of these networks during episodic retrieval. Together, findings from LV1 and LV2 of our T-PLS analysis indicate that older women and men at risk of AD engage similar brain regions during the encoding and retrieval of old vs. novel objects, and that these regions are comparable to episodic memory-related activity in cognitively healthy older adults.

### 4.2 Sex Differences in Task-related Brain Activity and Brain Activity-Behavior Correlations Suggesting Functional Dedifferentiation in Women

In addition to common task-related brain activation patterns in women and men during episodic encoding and retrieval (T-PLS LV1, LV2), our analyses identified generalized patterns of brain activity for encoding vs. retrieval of object-location associations in women, and more defined patterns distinguishing the encoding and retrieval of object-location associations vs. object-only information in men. In particular, T-PLS LV3 demonstrated no distinct activation patterns in women at encoding, and general activation of positive salience areas for object-location retrieval that were common to women and men. The B-PLS provided further evidence of generalized phase-related network activations in women, where memory performance – including source hits as well as failures – associated with activity in positive salience regions during encoding vs. negative salience regions at retrieval. In contrast, men exhibited distinct activation of positive salience regions for the encoding and retrieval of object-location associations, and of negative salience regions for object-only encoding and retrieval (-TPLS LV3). The B-PLS further identified a distinct network of retrieval-related brain activation in men, where associative retrieval (source hits) correlated with activity in positive salience regions, and object-only retrieval (source failures) correlated with activity in negative salience regions. These findings extend our previous reports of differences in brain activity and brain-behavior correlations during episodic encoding based on sex (Subramaniapillai et al., 2019) and genetic risk factors for AD (Rajah et al., 2017), highlighting sex differences in episodic encoding and retrieval related activity in older adults with family history of AD.

The activation patterns revealed in our T-PLS and B-PLS analyses may reflect a dedifferentiation of brain function (i.e., increasing interdependence among different brain processes and functions; S.-C. Li & Lindenberger, 1999; Rajah & D’Esposito, 2005) in older women with genetic risk of AD development. Although both sexes activated the medial, orbital and dorsal/superior PFC, caudate, middle temporal, occipital, and cingulate cortices for successful associative encoding, men additionally activated superior temporal, parietal, and dorsomedial frontal cortices. Moreover, whereas encoding related activity in the insula/ventrolateral PFC, posterior cingulate, and inferior parietal (i.e., supramarginal) cortex generally correlated with better subsequent performance – including source hits and failures – in women, these regions were distinctly active for subsequent object-only retrieval (source failures) in men. Similarly, whereas retrieval related activity in parahippocampal, anterior cingulate, precuneus, medial occipital, and ventrolateral PFC supported general memory performance in women, this activity was specific to object-only memory with source failure in men. These findings suggest that older at-risk women draw on generalized networks involving inferior and medial frontal, parietal, parahippocampal, and occipital cortex for episodic encoding and retrieval.

Dedifferentiation of brain function is widely reported with increasing age (Koen & Rugg, 2019). For example, studies have demonstrated age-related reductions in the functional specificity of parahippocampal and inferior temporal regions (Park et al., 2004) and episodic memory-related changes in the connections between the caudate and default-mode network (Rieckmann, Johnson, Sperling, Buckner, & Hedden, 2018). Previous research has further suggested that functional dedifferentiation associates with reduced performance on cognitive tasks (La Fleur, Meyer, & Dodson, 2018; Wilson, Segawa, Hizel, Boyle, & Bennett, 2012). Here we provide evidence for such dedifferentiation in women the absence of sex differences or deficits in episodic memory performance. Performance-independent dedifferentiation has also been revealed in healthy older adults during prospective memory tasks (Gonneaud et al., 2017). Dedifferentiation can therefore occur independently of performance changes, and may instead reflect a fundamental neurocognitive shift in approach to cognitive tasks. Our findings further suggest that such dedifferentiation may index a disparity in the way older women at risk of AD process episodic memory tasks, compared to their male counterparts.

Beyond a marker of age-related cognitive decline, dedifferentiation may reflect compensatory neurocognitive reorganization and/or pathological aging (S. C. Li, Lindenberger, & Sikstrom, 2001; Park & Reuter-Lorenz, 2009; Rajah & D’Esposito, 2005). Longitudinal studies in cognitively healthy older adults and AD converters suggest that cognitive dedifferentiation relates more strongly to terminal cognitive decline than to advancing age, and may be attributable to increasing neural pathology in late life stages (e.g., Batterham, Christensen, & Mackinnon, 2011; Hulur, Ram, Willis, Schaie, & Gerstorf, 2015; Wilson et al., 2012). Similarly, older adults in early stages of AD-related cognitive impairment exhibit generalized patterns of brain activity during associative memory task performance (Oedekoven, Jansen, Keidel, Kircher, & Leube, 2015) and rest (Bai et al., 2008), as well as memory-related hyperactivation of hippocampal and default mode regions (Dickerson et al., 2005; Nyberg, Andersson, Lundquist, Salami, & Wahlin, 2019). Resting and memory task-related hyperactivation of these regions also appears in young and older adults with increased genetic risk (i.e., family history or *APOE4* allele) of AD (Bookheimer et al., 2000; Filippini et al., 2009; Machulda et al., 2011; Quiroz et al., 2010) and may represent an early sign of pathological Aβ accumulation (Busche & Konnerth, 2016; Sperling et al., 2009). In the absence of performance deficits and presence of AD risk factors or biomarkers, such discrepant brain activity may serve as a compensatory mechanism (Cabeza et al., 2018; Han, Bangen, & Bondi, 2009). Findings from our B-PLS analysis support this compensatory interpretation of dedifferentiation, where phase (i.e., encoding and retrieval) related activity patterns supported general memory performance in at-risk women, whereas associative vs. object-only memory correlated with distinct retrieval related activations in at-risk men. Thus, the generalized activity patterns we found may represent an early biomarker for AD risk in older women. Neural dedifferentiation may therefore help in the development of clinical tools to better identify and intervene in early stages of dementia (Dodge & Marson, 2012), although potential changes in the degree of dedifferentiation with the progression of cognitive impairment (e.g., due to AD) remain unclear (Koen & Rugg, 2019).

Alongside dedifferentiation, the sex-specific activation patterns we observed support converging evidence suggesting that genetic (i.e., *+APOE4* or family history) risk for AD development is associated with greater encoding related activity in various regions of the default mode and dorsal attention networks, and that older women, compare to men, exhibit lower encoding-related temporal activity (McDonough, Festini, & Wood, 2020). However, our exploratory post-hoc analyses indicated no significant differences in event-related brain activity based on *APOE4* carrier status and a sex-by-*APOE4* effect only during the retrieval of object-only information (T-PLS LV3). Specifically, we found higher activation of negative salience regions, including ventral PFC, in *-APOE4* men compared to *-APOE4* women during object-only retrieval, with no significant sex differences in *+APOE4* participants. This pattern suggests that differences in object-only retrieval may appear less pronounced with elevated risk of AD. We found no other significant effects related to *APOE4*. This general absence of *APOE4*-specific effects is unsurprising given our null findings for *APOE* in past task fMRI analysis of this dataset and mixed effects of *APOE4* on episodic memory reported in the literature (McDonough et al., 2020), coupled with the small samples within each *APOE* genotype in this cohort. The sex-specific effects we found may therefore associate with the family history risk factor for AD, present in all participants of the PREVENT-AD cohort.

Our findings are among the first to identify sex differences in episodic memory-related brain activity and brain-behavior correlations in older adults with elevated genetic risk for AD. The evidence we present here is notable for several reasons. The PREVENT-AD cohort comprises a unique dataset of behavioral and task fMRI data in cognitively healthy older adults with first-degree (i.e., at least one parent or multiple sibling) history of AD. In addition, we carefully selected our female sample to match the available male sample based on age, years of education, and *APOE4* carrier status. Our resulting sample was further balanced in BMI, EYO, and other factors including raw task performance scores. Moreover, the present sample included both women and men with relatively high levels of education, a factor traditionally confounded with sex/gender effects on cognition (Angrisani, Lee, & Meijer, 2020; Tucker & Stern, 2011).

Historically, women have had different educational and occupational opportunities than men, and engaged in different lifestyle habits including levels of physical activity and participation in caregiver roles – factors that influence memory processes in normative aging and AD (Andrew & Tierney, 2018; Laws, Irvine, & Gale, 2018). The sex differences we observed in brain activation patterns are therefore unlikely to result from these factors, which are often confounded in studies of sex differences and of AD risk. Finally, given the greater number of older women compared to men diagnosed with AD (Beam et al., 2018), it is possible that our sample included more women, compared to men, at prodromal stages of AD. The identified dedifferentiation of brain function in women – i.e., failure of activation of a source-specific set of regions leading to object-only recognition – may therefore represent a sex-specific preclinical marker of AD-related functional neuropathology.

## 5. Limitations and Future Directions

In addition to limitations noted in our previous work (Rabipour et al., 2020), the number of men enrolled in PREVENT-AD limited our sample size. Analyzing a larger sample may have revealed different behavioral and brain activity patterns. Another caveat in the present analyses is the categorization of groups (i.e., men vs. women) based on a single, binary male/female self-report. Such limited questioning fails to capture possible nuances in sex and gender, including transsexual or transgender individuals whose self-identified sex/gender might not fit under the traditional binary categorization. Moreover, increasing evidence suggests a prominent impact of reproductive history and hormonal state (e.g., level of circulating estrogen, history of hormone replacement therapy, etc.) on episodic memory (Jacobs et al., 2016). Such information therefore represents an important consideration in any analysis of sex differences in memory function, and may contribute to episodic memory processes in healthy and pathological aging (Irvine et al., 2012); unfortunately, these data were incompletely available in the PREVENT-AD cohort. As the influence of sex on cognitive aging and AD risk becomes more apparent, studies are increasingly collecting these data to include as analytic variables

## 6. Conclusions

Here we examined sex differences in the relationship between memory performance and brain activity in age and education matched asymptomatic older adults with family history of AD. Despite largely comparable neuropsychological performance and task-related brain activation patterns during encoding and retrieval of old vs. novel objects in men and women, we found differences in brain activity and brain-behavior correlations. Notably, whereas women engaged sets of regions supporting general encoding vs. retrieval, men additionally engaged distinct regions supporting successful encoding and retrieval of object-location associations (source hits) vs. object-only information with source failure. Taken together, we conclude that older women and men with a family history of AD engage distinct activation patterns, which may reflect the use of different neurocognitive strategies to successfully encode and retrieve associative vs. object-only memory. Our future work will examine whether and how biological sex may influence these relationships longitudinally, including the potential role of brain structure and genotype in mediating these effects.

## Data Availability

Data from the PREVENT-AD study are available via https://openpreventad.loris.ca

## Acknowledgments

We thank members of the Rajah lab, the Cerebral Imaging Centre, and the StoP-AD Centre at the Douglas Hospital Research Centre for their helpful feedback on this work. We also acknowledge the Fonds de Recherche Québec – Santé, Natural Sciences and Engineering Research Council, and Canadian Institutes of Health Research for their support of our work.

PREVENT-AD was launched in 2011 as a $ 13.5 million, 7-year public-private partnership using funds provided by McGill University, the Fonds de Recherche du Québec – Santé, an unrestricted research grant from Pfizer Canada, the Levesque Foundation, the Douglas Hospital Research Centre and Foundation, the Government of Canada, and the Canada Fund for Innovation. Private sector contributions are facilitated by the Development Office of the McGill University Faculty of Medicine and by the Douglas Hospital Research Centre Foundation (http://www.douglas.qc.ca/).

## Author Contributions

Data creation was a collaborative effort from the PREVENT-AD Research Group. SR and MNR analyzed and interpreted the data with help from SR and SP. SR wrote the manuscript with contributions from MNR. All authors approved the final version of the manuscript.

